# PheWAS-ME: A web-app for interactive exploration of multimorbidity patterns in PheWAS

**DOI:** 10.1101/19009480

**Authors:** Nick Strayer, Jana K Shirey-Rice, Yu Shyr, Joshua C. Denny, Jill M. Pulley, Yaomin Xu

**Affiliations:** Department of Biostatistics, Vanderbilt University, Nashville, TN; Department of Biomedical informatics, Vanderbilt University, Nashville, TN; Center for Quantitative Sciences, Vanderbilt University Medical Center, Nashville, TN; Department of Medicine, Vanderbilt University School of Medicine, Nashville, TN; Office of Research, Vanderbilt University School of Medicine, Nashville, TN; Department of Medical Administration, Vanderbilt University School of Medicine, Nashville, TN

## Abstract

**Summary:** Electronic health records (EHRs) linked with a DNA biobank provide unprecedented opportunities for biomedical research in precision medicine. The Phenome-wide association study (PheWAS) is a widely-used technique for the evaluation of relationships between genetic variants and a large collection of clinical phenotypes recorded in EHRs. PheWAS analyses are typically presented as static tables and charts of summary statistics obtained from statistical tests of association between a genetic variant and individual phenotypes. Comorbidities are common and typically lead to complex, multivariate gene-disease association signals that are challenging to interpret. Discovering and interrogating multimorbidity patterns and their influence in PheWAS is difficult and time-consuming. We present PheWAS-ME: an interactive dashboard to visualize individual-level genotype and phenotype data side-by-side with PheWAS analysis results, allowing researchers to explore multimorbidity patterns and their associations with a genetic variant of interest. We expect this application to enrich PheWAS analyses by illuminating clinical multimorbidity patterns present in the data.

**Availability:** A demo PheWAS-ME application is publicly available at https://prod.tbilab.org/phewas_me/. Sample datasets are provided for exploration with the option to upload custom PheWAS results and corresponding individual-level data. The source code is available as an R package on GitHub (https://github.com/tbilab/multimorbidity_explorer).

## Introduction

Large-scale biobanks combined with electronic health records are increasingly available for clinical and translational research around the world (Chen et al., 2011; Cho et al., 2012; Gaziano et al., 2016; Investigators, 2019; McCarty et al., 2011; Sudlow et al., 2015). These data platforms typically provide subject-level information on a wide range of biomarkers along with detailed phenotype data and provide a highly anticipated paradigm shift for clinical and translational research in the era of precision medicine. The Phenome Wide Association Study is a statistical method to find associations across phenomes in the EHR with a given biomarker (e.g. SNPs). PheWAS quantifies associations between single SNP-phenotype pairs, which are blind to complex correlation structures present in phenotypes. When multiple phenotypes show a strong association with a genetic variant, researchers rely on domain expertise and more extensive interrogation of the data to determine potential causes. These include driver phenotypes (e.g., patients with a common disease taking a drug and then experiencing a common drug side effect), phenotype hierarchy, related diseases with an overlapping set of patients, or merely people with multiple diseases. Here we present PheWAS Multimorbidity Explorer (PheWAS-ME), a web application built using the programing language R and the Shiny library (Chang, Cheng, Allaire, Xie, & McPherson, 2018). PheWAS-ME allows researchers to interact with PheWAS results alongside the individual-level phenotype and genotype data that generated them. By visualizing individual-level data along with statistical results, the application provides a rich and explorable view into the patterns and relationships between phenotypes and the genotype being investigated. The interactive nature of the tool lets users enhance their interrogation of comorbidity patterns by delving into areas of interest on the phenome, such as a disease category, with custom visualizations. See Appendix B for a demonstration of the use of PheWAS-ME to parse the results of a PheWAS analysis to find novel phenotype associations.

## Implementation

Data needed to run PheWAS-ME are a standard PheWAS result table and the corresponding individual-level data. These results can be supplied to the app via a data loading screen or preloaded (see appendix C for full requirements). If desired, multiple comparisons correction can be performed on loaded data using either the Bonferroni (Dunn, 1961) or Benjamini-Hochberg (Benjamini & Hochberg 1995) methods.

After data are loaded, the app directs to the main visualization and analysis interface - an interactive dashboard including four views: SNP information, an interactive PheWAS Manhattan plot, multimorbidity UpSet plot, and a subject-phenotype bipartite network plot.

### Application state

PheWAS-ME works by filtering down to a list of ‘selected’ phenotypes. When a set of phenotypes is selected, the individual-level data are subset to just subjects who had one or more of the selected phenotypes in their records. This allows users to easily discard uninteresting or noisy phenotypes and focus in on potentially meaningful patterns using criteria like strength of the statistical association or phenotype category.

### SNP information panel

To provide context to the currently investigated SNP, the application provides a panel containing summary information (Figure 1A). Minor allele frequency in the provided subject population and the currently selected subset are shown as a bar chart. If the SNP of interest is present in an internal SNP annotation table sourced from dbSNP (Sherry et al., 2001) and VEP (McLaren et al., 2016), then additional information such as the minor allele, chromosome, and gene are provided.

**Figure 1.**
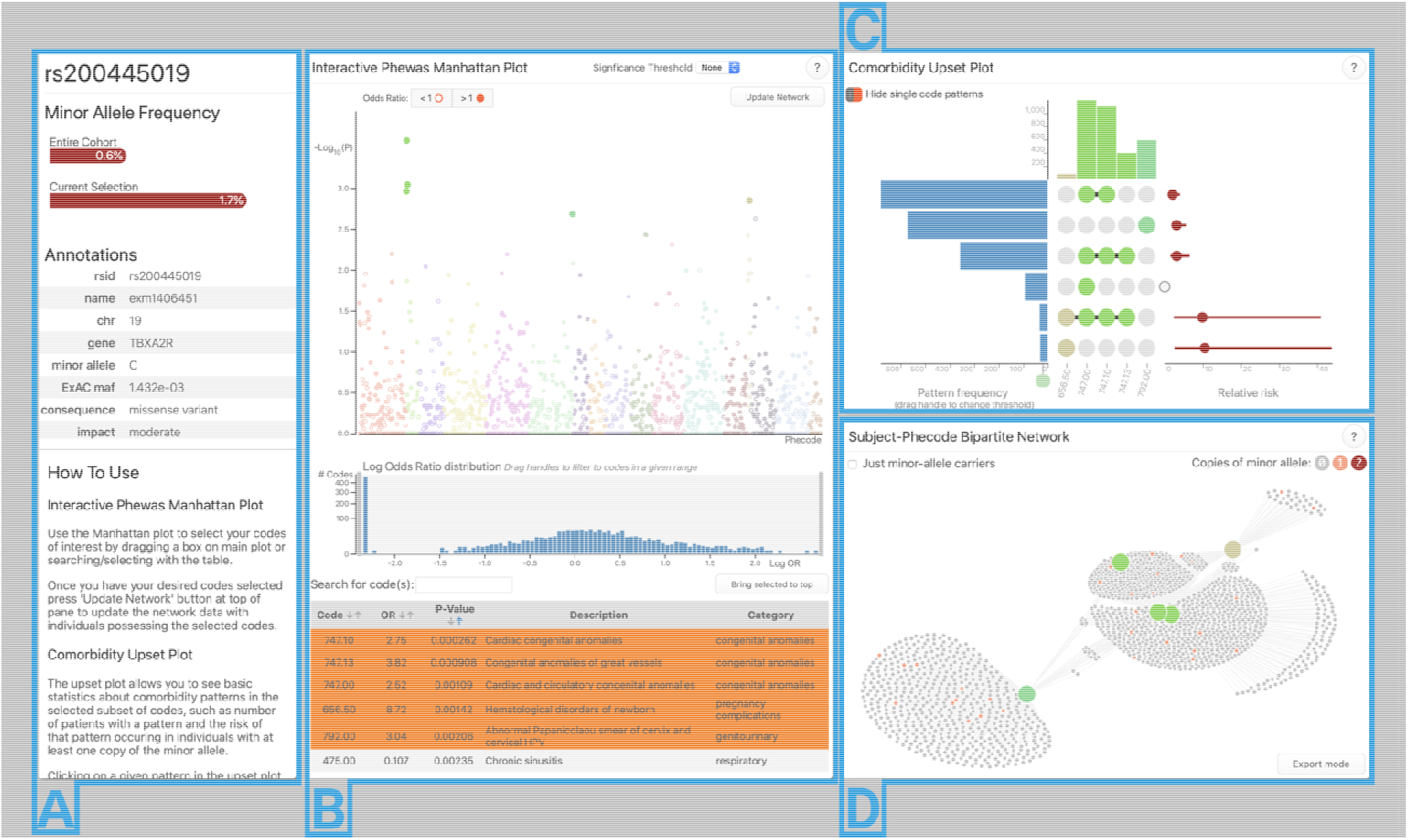

### Interactive PheWAS Manhattan plot

A manhattan plot (Figure 1B) is provided for the results of the PheWAS analysis (Denny et al., 2010). The x and y axis of this plot are phenotype diagnosis and statistical significance, respectively. Additional metadata from the supplied results table - such as name, description, and statistical results for a phenotype - are accessible by hovering over a phenotype’s point in the plot. Phenotypes can be selected for individual-level-data inspection by any combination of clicking, dragging a selection box, and searching in a table view below the plot. A histogram of the log-odds ratios for all phenotypes is provided and can be used to filter codes by ranges of association strength and direction.

### Multimorbidity UpSet plot

Figure 1C is an UpSet plot (Lex, Gehlenborg, Strobelt, Vuillemot, & Pfister, 2014). This plot shows the unique multimorbidity patterns seen in the individual-level data for the currently selected phenotypes as a matrix with columns as phenotypes and patterns (represented by filled phenotype columns) as rows. On the left side of the plot is a bar-chart displaying how many subjects had a multimorbidity pattern. To the right is a point estimate and 95% confidence interval of each pattern’s relative risk of occurring given that the subject has the given genetic variant of interest (calculated using Fisher’s Exact Test (Fisher, 1928)). When a pattern is selected, the subjects who have the pattern are highlighted in the network plot (Figure 1D). For more details on the upset plot we refer the reader to the original UpSet publication (Lex et al., 2014).

### Subject-Phenotype Bipartite Network

Individual-level data are visualized directly as a bipartite network. Phenotypes are represented as larger nodes (colored to match their point in the manhattan and upset plots) and subjects are represented as smaller nodes (colored by their number of copies of the SNP minor allele). A link is drawn between subjects and phenotypes if a subject was diagnosed with a phenotype. A physics-based layout simulation (Bostock, Ogievetsky, & Heer, 2011) is run in real-time as the data are filtered to position nodes with similar connection patterns close to each other. As the user investigates the network structure, phenotype nodes can be selected and isolated or removed from within the plot. An optional filtering mode limits the network to only subjects with one or more copies of the SNP’s minor allele, allowing investigation of genetics-driven patterns.

Greater detail of each section of PheWAS-ME is available via in-app help pages and the meToolkit package usage manual (see appendix C).

## Conclusion

In this paper we have provided a brief introduction to the application PheWAS Multimorbidity Explorer. This application takes PheWAS results and individual-level data, and enables researchers interactively explore complex multimorbidity patterns in PheWAS analyses.

## Data Availability

A demo PheWAS-ME application is publicly available at https://prod.tbilab.org/phewas_me/. Sample datasets are provided for exploration with the option to upload custom PheWAS results and corresponding individual-level data. The source code is available as an R package on GitHub (https://github.com/tbilab/meToolkit).

https://prod.tbilab.org/phewas_me_info/

